# Serum Magnesium, Prescribed Magnesium Replacement and Cardiovascular Events in Adults with Type 2 Diabetes: A National Cohort Study in U.S. Veterans

**DOI:** 10.1101/2025.05.19.25327928

**Authors:** Ying Yin, Yan Cheng, Andrew R. Zullo, Yijun Shao, Helen M. Sheriff, Charles Faselis, Simin Liu, Ali Ahmed, Qing Zeng-Treitler, Wen-Chih Wu

**Author notes:** **Correspondence:** Wen-Chih Wu, MD, MPH; Transforming Health Systems for Aging Veterans (THRIVE) Center of Innovation, Providence VA Healthcare System, Brown University Health Cardiovascular Institute, Alpert Medical School & School of Public Health, Brown University; Providence, RI. These authors contributed equally to this work and share senior authorship (Ali Ahmed, Qing Zeng-Treitler, Wen-Chih Wu).

## Abstract

**OBJECTIVE:** To investigate the relationship between serum magnesium levels, prescribed oral magnesium replacement, and major adverse cardiovascular events (MACE) in type-2 diabetes (T2D).

**RESEARCH DESIGN AND METHODS:** This national-wide retrospective study analyzed 1,284,940 US Veterans (≥18 years) with T2D who had outpatient serum magnesium testing between 1999-2021 in the Veterans Health Administration. The relationship between serum magnesium levels and MACE (hospitalizations for acute myocardial infarction, heart failure, or ischemic stroke, or all-cause mortality) was determined using multivariable-adjusted Cox-regression models. Using a new-user-design and propensity-score-matching analysis, we further related the use of prescribed oral magnesium and MACE among patients with hypomagnesemia (serum magnesium <1.8 mg/dL) and normomagnesemia (serum magnesium 1.8-2.3 mg/dl).

**RESULTS:** Of 1,284,940 patients with T2D, 229,210 (17.8%) patients had hypomagnesemia, and 117,674 (9.2%) patients had hypermagnesemia. Both hypomagnesemia and hypermagnesemia (serum magnesium >2.3 mg/dL) were linked to higher MACE risks (HRs 1.11-1.20 for hypo-and 1.04-1.39 for hypermagnesemia, respectively) compared to normomagnesemia. Oral magnesium was prescribed to 9.7% and 0.7% of patients with hypomagnesemia and normomagnesemia, respectively. After propensity-score-matching balanced across 64 baseline characteristics, oral magnesium was associated with a lower MACE risk in 40,766 matched patients with hypomagnesemia (HR 0.89; 95%CI, 0.84-0.93), especially those on proton-pump-inhibitors or thiazides. Oral magnesium was not related to MACE in 11,838 matched patients with normomagnesemia (HR 1.07; 95%CI, 0.97-1.17).

**CONCLUSIONS:** In patients with T2D, both hypomagnesemia and hypermagnesemia were associated with higher one-year MACE risks compared to normomagnesemia. Prescribed oral magnesium was associated with a reduced MACE risk in hypomagnesemia but not in normomagnesemia.

## 1. Introduction

Magnesium is an essential intracellular mineral involved in numerous enzymatic pathways. Normal serum levels range from 1.8–2.3 mg/dL, and around 13.5–47.7% of patients with type 2 diabetes experience hypomagnesemia.(1) Hypomagnesemia in type 2 diabetes patients can result from osmotic diuresis and loss of extracellular magnesium.(2–4) Additionally, commonly prescribed medications used in diabetic patients such as thiazide diuretics and proton pump inhibitors (PPIs) may exacerbate magnesium loss.(5)

Cardiovascular disease (CVD) is the leading cause of death among patients with type 2 diabetes. Hypomagnesemia is linked to oxidative stress,(6) inflammation,(7) endothelial dysfunction, and atherosclerosis,(8) further increasing CVD risk. Studies have not been conclusive on the relationship between serum magnesium levels and major adverse cardiovascular events (MACE: acute myocardial infarction [AMI], hospitalized heart failure [HF], ischemic stroke, and all-cause mortality) in patients with type 2 diabetes, partly due to limited sample size.(9, 10)

Meta-analyses of prospective cohort studies have indicated that increased dietary intake of magnesium-rich foods or the use of over-the-counter magnesium supplements is related to improvement in CVD risk factors(11–13) and risks of stroke(14)

(15) and HF.(16, 17) However, the majority of Americans have low magnesium intake.(18) Therefore, relying solely on the diet or supplements alone may be insufficient to maintain adequate body stores of magnesium, especially for patients with type 2 diabetes, who may need prescribed magnesium. There is a paucity of data relating prescribed oral magnesium in type 2 diabetes to MACE.

To address this gap, we leveraged the comprehensive electronic medical record (EMR) system from the Veterans Health Administration (VHA) and conducted a retrospective analysis of a nationwide cohort of Veterans with type 2 diabetes and serum magnesium levels. This study assesses the relationship between serum magnesium levels, prescribed oral magnesium for replacement, and MACE. We hypothesized that low serum magnesium levels are linked to adverse MACE outcomes, and prescribed oral magnesium may benefit those with hypomagnesemia.

## 2. Methods

### 2.1 Study Design

We conducted a retrospective study of a nationwide cohort of Veterans ≥18 years old diagnosed with type 2 diabetes using cross-linked EMR data from January 1, 1999, to December 31, 2022, within the VHA’s Corporate Data Warehouse (CDW), which includes comprehensive data on hospitalizations, clinic visits, laboratory results, and medications, and the VA-Medicare datasets to assess for healthcare utilization outside of the VHA for Medicare-eligible Veterans. We first evaluated the relationship between serum magnesium levels and MACE outcomes. We then examined the association between prescribed magnesium and MACE using a new-user design in patients with normal or low serum magnesium levels. The study was approved by the VA Institutional Review Board, Washington, DC, VA Medical Center.

### 2.2 Eligibility Criteria

Patients with type 2 diabetes were identified by two type 2 diabetes International Classification of Diseases (ICD) codes (**eTables 1**), one ICD code and one type 2 diabetes medication (**eTables 2**), or one ICD code and hemoglobin A1c ≥6.5%, based on the VHA’s “Diabetes Epidemiology Cohorts” criteria.(19) We identified 3,448,558 Veterans who had a new diagnosis of type 2 diabetes and received care within VHA between 1999 and 2021. We excluded 14 patients who were <18 years of age, 978,176 patients without at least one-year history of VHA care, and 1,185,428 patients without a serum magnesium test for a final study sample of 1,284,940 patients to relate serum magnesium levels to MACE. The date of the ambulatory magnesium test was used as the index date, henceforward.

To relate prescribed oral magnesium and MACE, we excluded patients who received prescribed oral and intravenous magnesium in the year before serum magnesium test (n=27,213) to avoid prevalent user bias, patients who had hypermagnesemia (serum magnesium >2.3 mg/dL) as they would not be eligible to receive magnesium replacement (n=115,619), and those who experienced MACE within 15 days of magnesium test (n=32,572) to provide a 15-day blanking period allowing time for magnesium prescription (**eFigure 1**). Thus, this analysis was restricted to 211,128 patients with hypomagnesemia (serum magnesium <1.8 mg/dL) and 893,768 patients with normomagnesemia (serum magnesium 1.8-2.3 mg/dL).

### 2.3 Treatment Strategies

Given that oral replenishment of the body’s magnesium stores takes time,(20) patients from our initial cohort eligible for prescribed magnesium were allocated to the treatment group if they received a ≥30-day prescription for oral magnesium within 15 days following a serum magnesium test (n=4,466 excluded). Data on magnesium prescriptions (including magnesium oxide, magnesium gluconate, magnesium chloride, and magnesium sulfate) were obtained from VHA pharmacy data. Magnesium citrate and magnesium hydroxide were excluded, as they were primarily used for bowel regimens. As we are only interested in oral magnesium, patients with intravenous magnesium prescriptions within 15 days were also excluded (n = 174). All analyses focused on an intention-to-treat estimand – the effect of initiating magnesium versus not, regardless of subsequent treatment discontinuation or switching among treatment groups.

### 2.4 Outcomes

The <u>primary</u> outcome was MACE: hospitalizations with AMI, HF, ischemic stroke, or all-cause mortality within one year of the index date, with the last date of outcome accrual being December 31, 2022. Data on hospitalizations were abstracted using ICD codes (eTable 1) from VHA hospital records and Medicare claims for non-VHA facilities. The <u>secondary</u> outcome was all-cause mortality. Patients’ vital status and death date were obtained from the VHA CDW, which compiled records from death certificates, the Social Security Administration Death Master File, and the Centers for Medicare & Medicaid Services Vital Status Files.

### 2.5 Covariates

Selected baseline conditions were defined using ICD codes (**eTable 1)** documented at any time before the index date. Baseline Gagne comorbidity scores(21), a weighted score to assess overall comorbidity, were calculated. Medications (**eTable 2**) were identified from outpatient pharmacy data within one year before the index date. We also gathered the most recent data for vital signs and laboratory values within one year prior to the index date. The calendar year of the index date, patients’ geographic location by Veterans-Integrated-Services-Networks (VISNs), and the rurality of the residence were included for a total of 64 covariates extracted (**eFigure 2**).

Missing values were coded as not present for categorical variables. For continuous variables (**eTable 3**), missing values were imputed by single imputation with a fitted general linear model on age, sex, race and ethnicity. Due to the large sample size, absolute standardized differences (ASD) were calculated to compare baseline covariates between the analyzed groups.

### 2.6 Statistical analysis

#### 2.6.1 Serum Magnesium Level and MACE outcome

We performed multivariable Cox regression analysis adjusted for 64 baseline covariates to assess the association between serum magnesium levels and one-year MACE outcomes.

#### 2.6.2 Propensity Score Matching

To estimate the association between prescribed magnesium and outcomes, we used propensity scores (PS) to assemble balanced cohorts of prescribed magnesium versus no prescribed magnesium in an outcome-blinded manner, mimicking a randomized controlled trial (RCT).(22, 23) To analyze patients with <u>hypomagnesemia</u>, we used a non-parsimonious multivariable logistic regression model to calculate PS for the receipt of prescribed magnesium for each of the 211,128 patients based on the 64 baseline characteristics. We used a greedy matching protocol to match patients with and without prescribed magnesium based on PS to five, four, three, two, and one decimal places in five steps. This yielded 20,383 patients in each group, forming a matched cohort of 40,766 patients (**eFigure 1**). The balance in baseline characteristics was measured using ASD in the post-match cohort.(24) An ASD <10% is considered inconsequential. For patients with <u>normomagnesemia</u>, we repeated the above approach and assembled a PS-matched cohort of 11,838 patients with 1:1 matching (**eFigure 1**).

The Kaplan-Meier method was used to estimate the cumulative probabilities of MACE and death. After confirming the assumption of risk proportionality by visualizing the Schoenfeld residuals, we used Cox regression models to estimate hazard ratios (HRs) and 95% confidence interval (95%CI) for the outcomes.(25, 26)

#### 2.6.3 Sensitivity Analysis

To assess the robustness of the effect estimates using an alternative analytic methodology, we also determined the association between prescribed magnesium and outcomes using multivariable Cox regression models adjusted for all 64 covariates in the pre-PS-matched cohort for the hypomagnesemia and the normomagnesemia groups, respectively, and estimated adjusted HRs (95%CI).

#### 2.6.4 Subgroup analyses

Subgroup analyses were performed to evaluate whether the association between prescribed magnesium and outcomes varied across participant characteristics (i.e., effect measure modification). Subgroups were based on demographics (sex and race), serum magnesium levels, comorbidity (HF), medications that can lead to hypomagnesemia (diuretics, PPIs, metformin) or interacts with magnesium (vitamin D) to determine for heterogeneity of associations, if any, between prescribed magnesium and MACE, in the PS-matched cohort for both hypomagnesemia and normomagnesemia groups.

For each subgroup variable, we fit separate PS within each subgroup stratum, and individuals were re-matched to ensure a covariate balance between treatment groups within subgroups, confirmed using ASD < 0.10.(27) After PS-matching, Cox proportional hazards regression was employed to estimate the treatment effects on MACE outcome within each subgroup. We then aggregated data from the propensity-score-balanced strata into a single dataset and fit regression models, including interaction terms between prescribed magnesium and each subgroup variable (i.e., each in a separate model), to estimate p-values for homogeneity. A p-value below 0.05 indicated a statistically significant difference in the estimated effect across subgroup strata and potential effect modification by that subgroup variable.

#### 2.6.5 Software

Statistical analyses were conducted using Python. The graphics were generated using the lifeline, sklearn, scipy, and matplotlib packages.

## 3. Results

### 3.1 Serum Magnesium and MACE in Type-2 Diabetes

Of the 1,284,940 patients with ambulatory serum magnesium levels after type 2 diabetes diagnosis, 229,210 (17.8%) patients had hypomagnesemia, and 117,674 (9.2%) patients had hypermagnesemia. Both hyper-and hypomagnesemia patients had a higher comorbidity burden measured by Gagne comorbidity score compared to patients with normomagnesemia. Patients with hypomagnesemia had a longer diabetes duration and a higher prevalence of hypertension, anemia, alcohol abuse, liver disease, and metformin use. Conversely, hypermagnesemia patients were more likely to have a history of HF, respiratory failure, or reduced kidney function (**Table 1**).

**Table 1:**
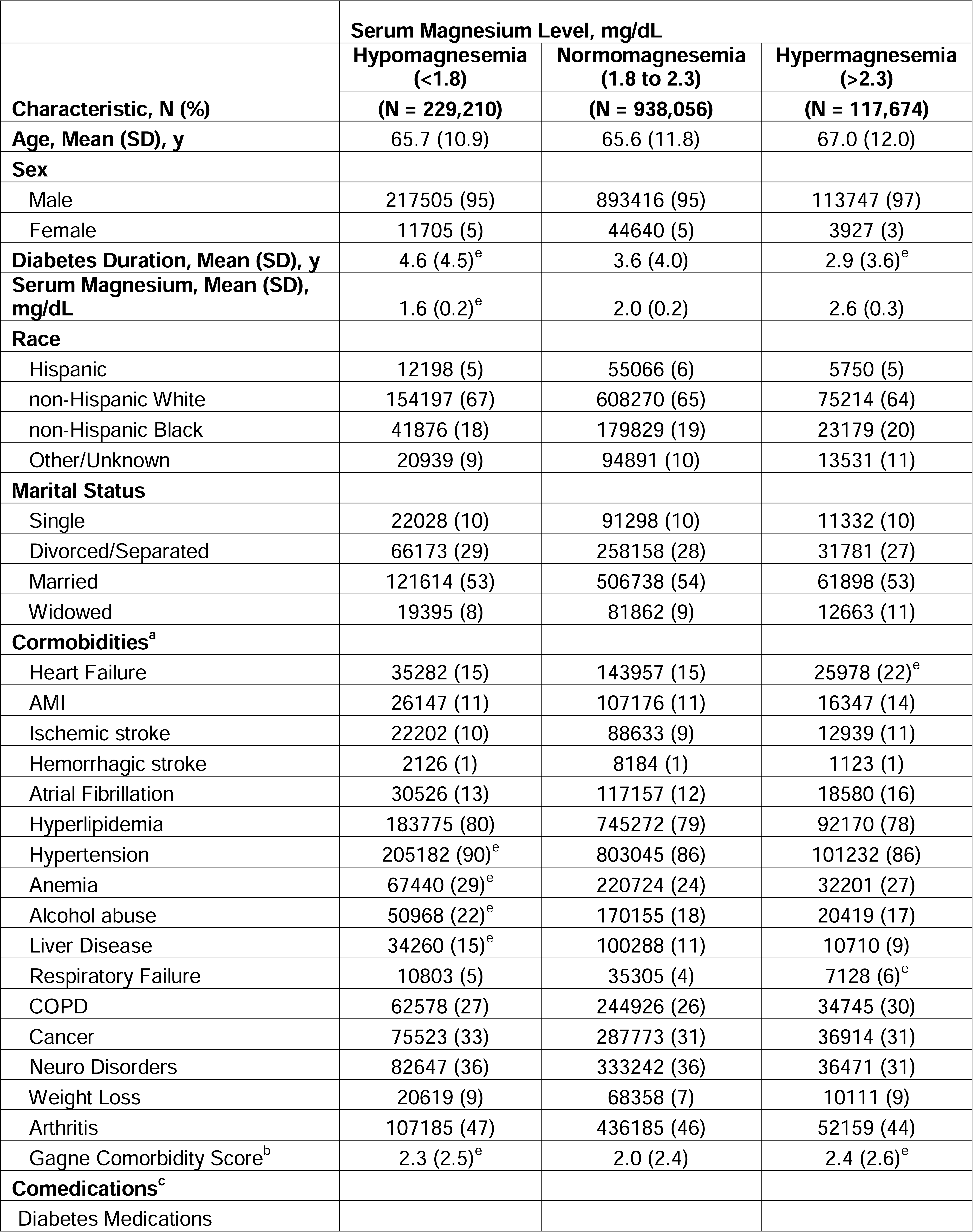
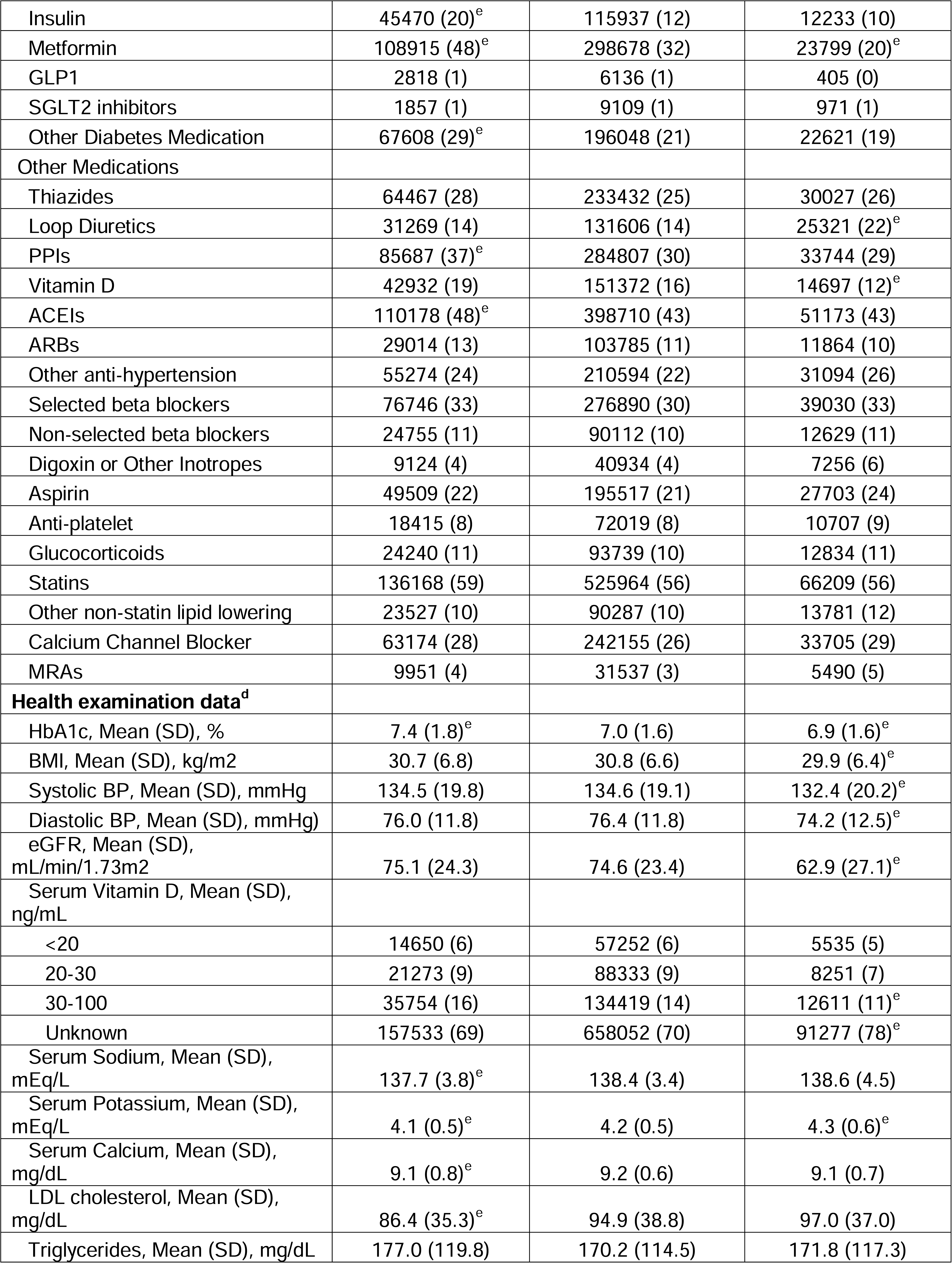
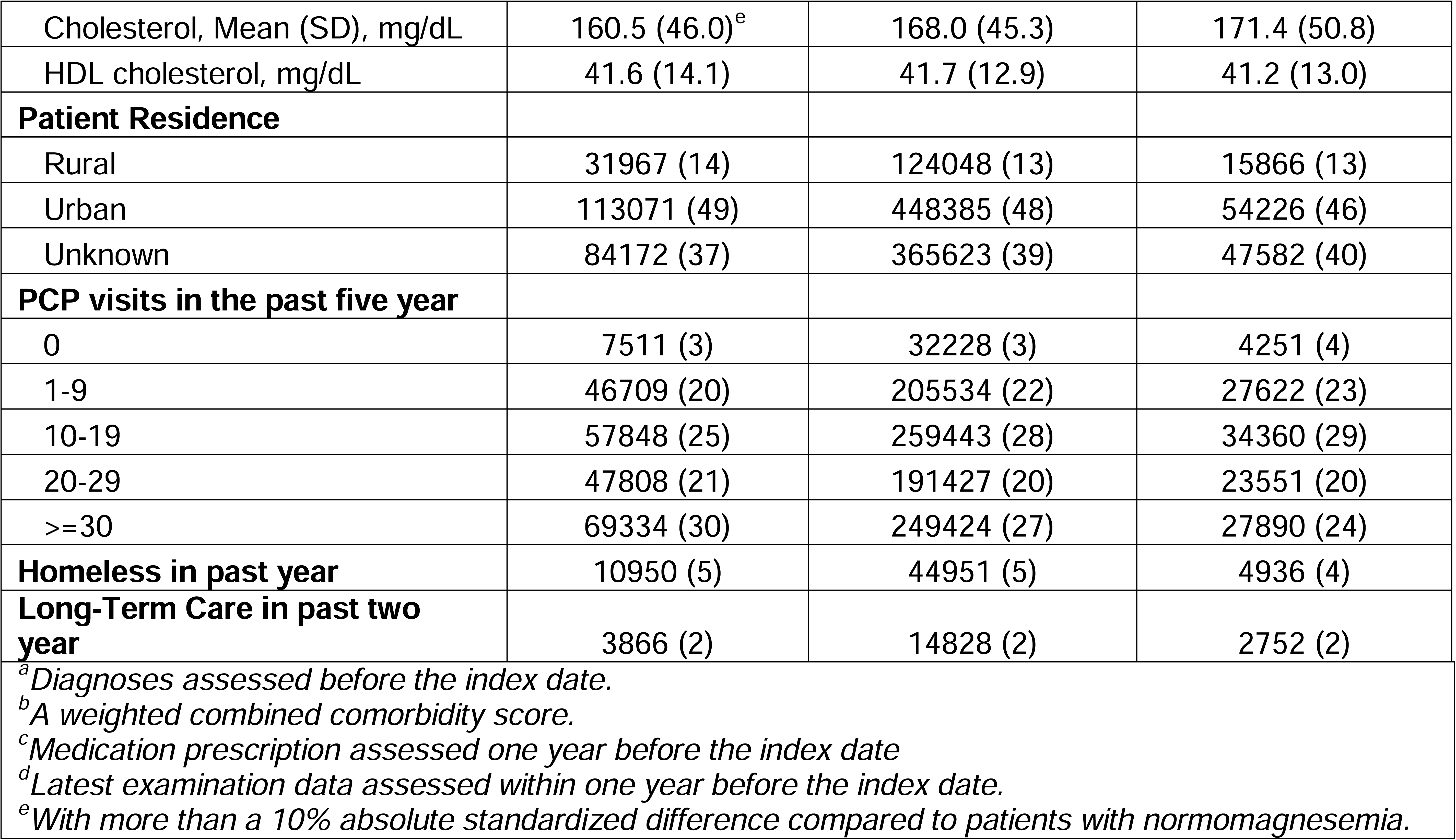
Baseline Characteristics of Patients with Type 2 Diabetes Categorized by Three Serum Magnesium Levels.

Compared to patients with normomagnesemia, patients in either hypomagnesemia or hypermagnesemia groups were associated with elevated hazards for MACE. The hazards increased with the severity of the serum magnesium abnormality in both high and low magnesium levels in a parabolic pattern. Specifically, patients with serum magnesium levels <1.4 mg/dL had a 20% higher, and those with levels >2.9 mg/dL had a 39% higher risk of MACE at one year, compared with the normomagnesemia group (Figure 1).

**Figure 1:**
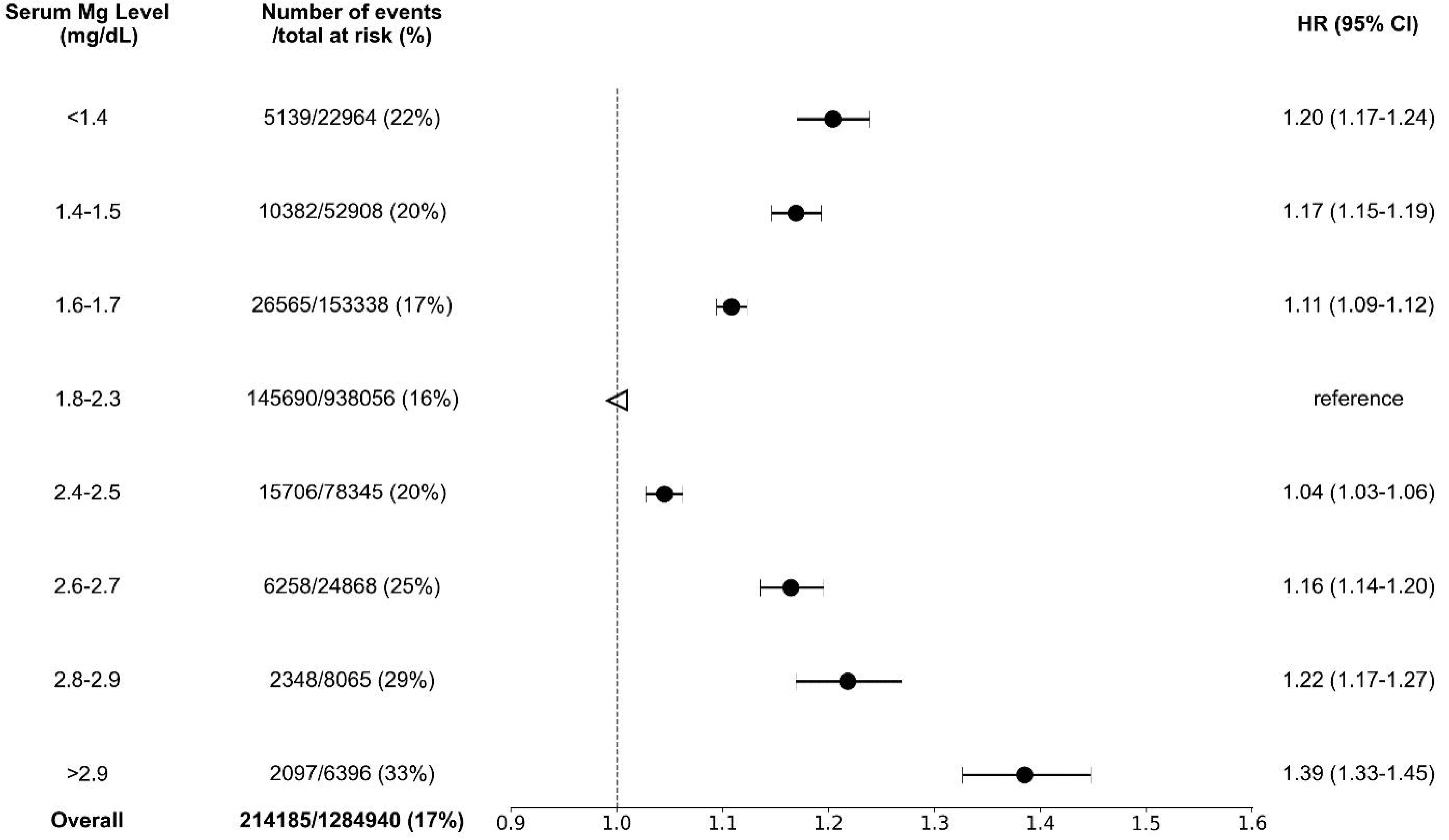
Adjusted Hazard Ratios for Baseline Magnesium Levels and Time to MACE in Patients with Type 2 Diabetes The dots with error bars represent the adjusted hazard ratios (95% confidence intervals) for time to MACE within one year of an ambulatory magnesium test in patients with type 2 diabetes and varying serum magnesium levels, compared with patients with type 2 diabetes and normal serum magnesium levels. The analysis used a multivariate Cox regression model adjusted for 64 variables, including demographics, comorbidities, medication history, and recent lab tests.

### 3.2 Prescribed Oral Magnesium and Outcomes

Prescribed oral magnesium was observed in 9.7% of patients with hypomagnesemia and 0.7% of patients with normomagnesemia. The median duration of magnesium prescription was 90 days (interquartile range [IQR]: 30-176 days), with a median dosage of 420 mg/day (range: 140-500 mg/day). Approximately 99% of the prescriptions were for magnesium oxide.

#### 3.2.1 Oral Magnesium in Hypomagnesemia

Compared to patients without magnesium prescription, patients with prescribed magnesium were more likely to be of White race, have a longer diabetes duration, and have a higher prevalence of hyperlipidemia, hypertension, and treatment with metformin, PPIs, vitamin D and statins (**Table 2**). After PS-matching, n=20,383 patients with prescribed magnesium were PS-matched to n=20,383 patients without magnesium prescription with adequate balance on 64 baseline covariates (**eFigure 2A**). The patients in the PS-matched cohort had a mean (SD) age of 66 (10) years and were comprised of 5% women and 72% White, with a mean (SD) serum magnesium level of 1.5 (0.2) mg/dL and an estimated glomerular filtration rate (eGFR) of 74.1 (22.5) mL/min/1.73 m^2^.

**Table 2.** Baseline Characteristics of Patients with Type 2 Diabetes and Hypomagnesemia Initiated on Prescribed Oral Magnesium, Before and After Propensity Score Matching.

| Characteristic, N (%) | Before matching<br>(n=211,128) |  | After matching<br>(n=40,766) |  |
| --- | --- | --- | --- | --- |
|  | Prescribed Magnesium |  | Prescribed Magnesium |  |
|  | No | Yes | No | Yes |
|  | (n= 190,683) | (n= 20,445) | (n= 20,383) | (n= 20,383) |
| **Age, Mean (SD), y** | 65.6 (10.9) | 66.5 (10.1) | 66.6 (10.3) | 66.5 (10.1) |
| **Sex** |  |  |  |  |
| Male | 180965 (95) | 19372 (95) | 19339 (95) | 19311 (95) |
| Female | 9718 (5) | 1073 (5) | 1044 (5) | 1072 (5) |
| **Diabetes Duration, Mean (SD), y** | 4.5 (4.5) | 5.5 (4.8)<sup>e</sup> | 5.6 (4.8) | 5.5 (4.8) |
| **Serum Magnesium, Mean (SD), mg/dL** | 1.6 (0.2) | 1.5 (0.2)<sup>e</sup> | 1.5 (0.2) | 1.5 (0.2) |
| **Race** |  |  |  |  |
| Hispanic | 10559 (6) | 772 (4) | 713 (3) | 770 (4) |
| non-Hispanic White | 127468 (67) | 14812 (72)<sup>e</sup> | 14866 (73) | 14760 (72) |
| non-Hispanic Black | 35208 (18) | 3155 (15) | 3059 (15) | 3150 (15) |
| Other/Unknown | 17448 (9) | 1706 (8) | 1745 (9) | 1703 (8) |
| **Marital Status** |  |  |  |  |
| Single | 18498 (10) | 1701 (8) | 1701 (8) | 1697 (8) |
| Divorced/Separated | 55079 (29) | 5578 (27) | 5509 (27) | 5561 (27) |
| Married | 101041 (53) | 11525 (56) | 11504 (56) | 11492 (56) |
| Widowed | 16065 (8) | 1641 (8) | 1669 (8) | 1633 (8) |
| **Cormobidities<sup>a</sup>** |  |  |  |  |
| Heart Failure | 26785 (14) | 3028 (15) | 3074 (15) | 3019 (15) |
| AMI | 20489 (11) | 2091 (10) | 2079 (10) | 2084 (10) |
| Ischemic stroke | 17691 (9) | 1734 (8) | 1756 (9) | 1730 (8) |
| Hemorrhagic stroke | 1659 (1) | 145 (1) | 149 (1) | 145 (1) |
| Atrial Fibrillation | 23787 (12) | 2888 (14) | 2925 (14) | 2873 (14) |
| Hyperlipidemia | 152162 (80) | 17385 (85)<sup>e</sup> | 17286 (85) | 17327 (85) |
| Hypertension | 169799 (89) | 18882 (92)<sup>e</sup> | 18752 (92) | 18823 (92) |
| Anemia | 54195 (28) | 6087 (30) | 6069 (30) | 6063 (30) |
| Alcohol abuse | 41360 (22) | 4497 (22) | 4516 (22) | 4477 (22) |
| Liver Disease | 27556 (14) | 3029 (15) | 2888 (14) | 3023 (15) |
| Respiratory Failure | 8394 (4) | 590 (3) | 600 (3) | 590 (3) |
| COPD | 50423 (26) | 5599 (27) | 5539 (27) | 5579 (27) |
| Cancer | 62495 (33) | 6359 (31) | 6280 (31) | 6339 (31) |
| Neuro Disorders | 67338 (35) | 7935 (39) | 7833 (38) | 7909 (39) |
| Weight Loss | 16505 (9) | 1562 (8) | 1603 (8) | 1557 (8) |
| Arthritis | 87972 (46) | 10249 (50) | 10144 (50) | 10208 (50) |
| Gagne Comorbidity Score<sup>b</sup> | 2.2 (2.5) | 2.2 (2.4) | 2.2 (2.4) | 2.2 (2.4) |
| **Comedications<sup>c</sup>** |  |  |  |  |
| Diabetes Medications |  |  |  |  |
| Insulin | 36989 (19) | 4719 (23) | 4648 (23) | 4713 (23) |
| Metformin | 89716 (47) | 11753 (57)<sup>e</sup> | 11704 (57) | 11712 (57) |
| GLP1 | 2287 (1) | 327 (2) | 322 (2) | 327 (2) |
| SGLT2 inhibitors | 1504 (1) | 207 (1) | 194 (1) | 206 (1) |
| Other Diabetes Medication | 55719 (29) | 7065 (35)<sup>e</sup> | 7051 (35) | 7045 (35) |
| Other Medications |  |  |  |  |
| Thiazides | 53071 (28) | 6531 (32) | 6459 (32) | 6502 (32) |
| Loop Diuretics | 23632 (12) | 3147 (15) | 3174 (16) | 3136 (15) |
| PPIs | 68107 (36) | 9151 (45)<sup>e</sup> | 8945 (44) | 9100 (45) |
| Vitamin D | 33566 (18) | 4828 (24)<sup>e</sup> | 4796 (24) | 4811 (24) |
| ACEIs | 90817 (48) | 10466 (51) | 10397 (51) | 10433 (51) |
| ARBs | 23504 (12) | 3134 (15) | 3125 (15) | 3117 (15) |
| Other anti-hypertension | 44324 (23) | 5310 (26) | 5231 (26) | 5289 (26) |
| Selected beta blockers | 62014 (33) | 7251 (35) | 7226 (35) | 7231 (35) |
| Non-selected beta blockers | 19443 (10) | 2458 (12) | 2429 (12) | 2445 (12) |
| Digoxin or Other Inotropes | 7016 (4) | 884 (4) | 920 (5) | 879 (4) |
| Aspirin | 40048 (21) | 4405 (22) | 4351 (21) | 4386 (22) |
| Anti-platelet | 14750 (8) | 1684 (8) | 1642 (8) | 1676 (8) |
| Glucocorticoids | 19131 (10) | 1992 (10) | 1917 (9) | 1985 (10) |
| Statins | 111767 (59) | 13462 (66)<sup>e</sup> | 13448 (66) | 13411 (66) |
| Other non-statin lipid lowering | 19304 (10) | 2292 (11) | 2267 (11) | 2279 (11) |
| Calcium Channel Blocker | 51103 (27) | 6198 (30) | 6115 (30) | 6172 (30) |
| MRAs | 7251 (4) | 1086 (5) | 1068 (5) | 1081 (5) |
| **Health examination data<sup>d</sup>** |  |  |  |  |
| HbA1c, Mean (SD), % | 7.4 (1.8) | 7.4 (1.6) | 7.3 (1.6) | 7.4 (1.6) |
| BMI, Mean (SD), kg/m2 | 30.7 (6.8) | 31.3 (6.7) | 31.3 (6.9) | 31.3 (6.8) |
| Systolic BP, Mean (SD), mmHg | 134.8 (19.8) | 133.4 (18.8) | 133.3 (19.1) | 133.4 (18.8) |
| Diastolic BP, Mean (SD), mmHg | 76.0 (11.8) | 75.7 (11.3) | 75.5 (11.3) | 75.7 (11.3) |
| eGFR, Mean (SD), mL/min/1.73m2 | 75.5 (24.4) | 74.2 (22.2) | 74.0 (23.3) | 74.2 (22.2) |
| Serum Vitamin D, Mean (SD), ng/mL |  |  |  |  |
| <20 | 11780 (6) | 1788 (9)<sup>e</sup> | 1804 (9) | 1784 (9) |
| 20-30 | 17328 (9) | 2499 (12)<sup>e</sup> | 2494 (12) | 2492 (12) |
| 30-100 | 28700 (15) | 4413 (22)<sup>e</sup> | 4453 (22) | 4389 (22) |
| Unknown | 132875 (70) | 11745 (57)<sup>e</sup> | 11632 (57) | 11718 (57) |
| Serum Sodium, Mean (SD), mEq/L | 137.7 (3.7) | 138.1 (3.5)<sup>e</sup> | 138.1 (3.6) | 138.1 (3.5) |
| Serum Potassium, Mean (SD), mEq/L | 4.1 (0.5) | 4.1 (0.5) | 4.1 (0.5) | 4.1 (0.5) |
| Serum Calcium, Mean (SD), mg/dL | 9.1 (0.8) | 9.2 (0.7)<sup>e</sup> | 9.2 (0.8) | 9.2 (0.7) |
| LDL cholesterol, Mean (SD), mg/dL | 87.0 (35.3) | 82.2 (34.0)<sup>e</sup> | 81.8 (33.9) | 82.2 (34.1) |
| Triglycerides, Mean (SD), mg/dL | 177.6 (120.1) | 178.8 (121.3) | 177.9 (119.5) | 178.8 (121.3) |
| Cholesterol, Mean (SD), mg/dL | 161.2 (46.0) | 155.6 (43.7)<sup>e</sup> | 154.9 (41.1) | 155.6 (43.7) |
| HDL cholesterol, mg/dL | 41.5 (13.8) | 42.3 (15.2) | 42.5 (15.0) | 42.3 (15.2) |
| **Patient Residence** |  |  |  |  |
| Rural | 26004 (14) | 3283 (16) | 3291 (16) | 3264 (16) |
| Urban | 93229 (49) | 10263 (50) | 10284 (50) | 10232 (50) |
| Unknown | 71450 (37) | 6899 (34) | 6808 (33) | 6887 (34) |
| **PCP visits in the past five year** |  |  |  |  |
| 0 | 6542 (3) | 542 (3) | 545 (3) | 540 (3) |
| 1-9 | 40173 (21) | 3394 (17)<sup>e</sup> | 3384 (17) | 3388 (17) |
| 10-19 | 48662 (26) | 4946 (24) | 5105 (25) | 4932 (24) |
| 20-29 | 39645 (21) | 4481 (22) | 4341 (21) | 4462 (22) |
| >=30 | 55661 (29) | 7082 (35)<sup>e</sup> | 7008 (34) | 7061 (35) |
| **Homeless in past year** | 9112 (5) | 807 (4) | 831 (4) | 806 (4) |
| **Long-Term Care in past two year** | 3080 (2) | 172 (1) | 181 (1) | 172 (1) |

<sup>a</sup>Diagnoses assessed before the index date.
<sup>b</sup>A weighted combined comorbidity score.
<sup>c</sup>Medication prescription assessed one year before the index date
<sup>d</sup>Latest examination data assessed within one year before the index date
<sup>e</sup>With more than a 10% absolute standardized difference compared to patients with no prescribed magnesium before propensity score matching

In these PS-matched patients, MACE occurred in 13.6% of the prescribed magnesium and 15.1% of the non-magnesium groups (**eTable 5**). Cox-regression showed that prescribed magnesium was associated with decreased one-year risks of MACE, HR 0.89 (95%CI 0.84–0.93) (**Figure 2A**) and death (HR: 0.86 (95% CI, 0.80-0.93) (**eFigure 3A**). Sensitivity analyses using multivariate Cox regression modeling applied to the pre-PS-matched cohort of patients yielded similar results. The adjusted HRs associated with prescribed magnesium compared to non-magnesium were 0.93 (95% CI, 0.89-0.96) for MACE and 0.93 (95% CI, 0.88-0.99) for all-cause mortality (**eTable 5**).

**Figure 2:**
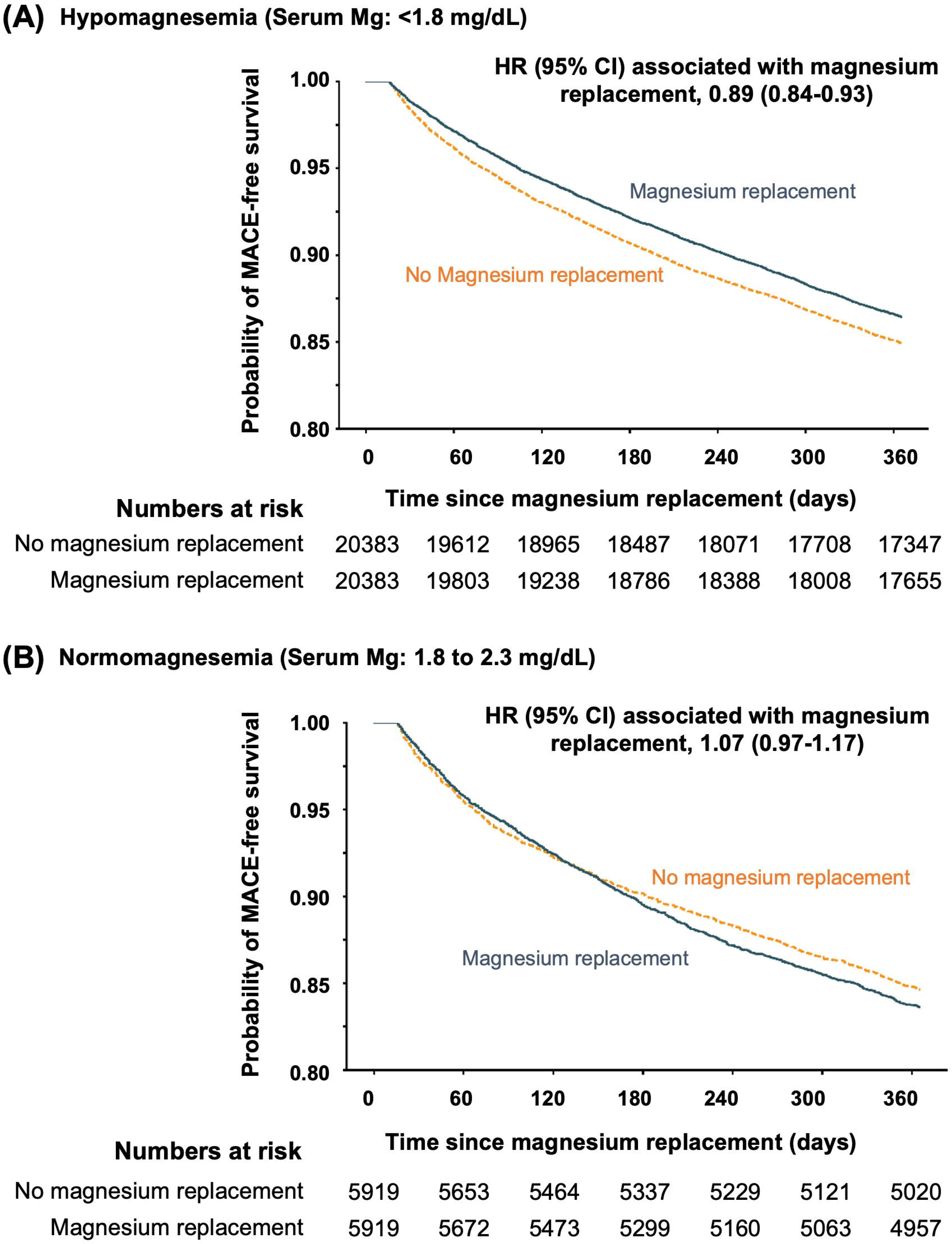
One-Year Kaplan-Meier Curves for MACE-Free Survival by Initiation of Prescribed Magnesium in Propensity Score-Matched Cohorts of Patients with Type 2 Diabetes and A) Hypomagnesemia, or B) Normomagnesemia

The association between prescribed magnesium and MACE in the PS-matched cohort was homogeneous across subgroups of age, sex, race, serum magnesium, HF, and certain medications (metformin, loop diuretics, vitamin D). Exceptions were noted for subgroups stratified by the use of PPIs (p = 0.026 for interaction) or thiazides (p = 0.039 for interaction), where the association of prescribed magnesium with lower risks of MACE was more substantial for those on PPIs or thiazides compared to those not on the respective medication(s) (**eFigure 4**).

#### 3.2.2 Oral Magnesium in Normomagnesemia

Compared to patients without magnesium prescription, those in the prescribed magnesium group had a higher Gagne comorbidity score and a higher prevalence of a history of HF, atrial fibrillation, anemia, alcohol abuse, liver disease, and neurologic disorders. They were also more likely to be treated with loop diuretics, non-selective beta-blockers, mineralocorticoid-receptor-antagonists, and PPIs (**eTable 4**). After PS-matching, n=5,919 patients with prescribed magnesium were PS-matched to n=5,919 patients without magnesium prescription (**eFigure 2B**). The patients in the PS-matched cohort had an average (SD) age of 65 (11) years, were comprised of 6% women and 68% White, and had a mean (SD) serum magnesium level of 2.0 (0.2) mg/d and an eGFR of 73.9 (24.5) mL/min/1.73 m^2^.

In the PS-matched cohort of the normomagnesemic group, MACE occurred in 16.4% of patients who received prescribed magnesium compared to 15.4% in the non-magnesium group (**eTable 5**). Prescribed magnesium was not significantly associated with risks of MACE [HR 1.07 (95% CI, 0.97–1.17)] (**Figure 2B),** or death [HR 1.00 (95% CI, 0.87–1.15)] (**eFigure 3B**). Subgroup analyses showed the association between prescribed magnesium and MACE in the normomagnesemic PS-matched cohort to be homogeneous, except for subgroups stratified by the use of PPIs (p = 0.007 for interaction), which showed a higher risk of MACE in patients with prescribed magnesium and not on PPIs (**eFigure 5**).

Sensitivity analyses using multivariate-Cox-regression modeling in the pre-PS-matched cohort showed that prescribed magnesium in normomagnesemic patients was associated with higher risks of MACE, HR 1.14 (95%CI 1.07-1.21), and death, HR 1.11 (95%CI 1.01-1.23).

## 4. Discussion

In a nationwide cohort of over 1.2 million Veterans with type 2 diabetes, 18% had hypomagnesemia, and 9% had hypermagnesemia. Both hypomagnesemia and hypermagnesemia were associated with higher risks of one-year MACE and death compared to patients with normomagnesemia. In patients with hypomagnesemia, prescribed magnesium was associated with reduced one-year risks of MACE and death, whereas in patients with normomagnesemia, prescribed magnesium was not associated with MACE.

### 4.1 Hypomagnesemia and Cardiovascular Outcomes

Previous research on the relationship between low serum magnesium levels and increased mortality have focused on critically ill inpatients or those with end-stage renal disease.(28–32) Observations from outpatient cohorts with type 2 diabetes have been inconsistent in relating serum magnesium levels to MACE, likely due to limited sample sizes. A cross-sectional study (n = 450) of patients with type 2 diabetes demonstrated a lower incidence of coronary heart disease with higher serum magnesium levels,(33) while the Fremantle Diabetes Study (n = 940) identified an inversely association between serum magnesium and stroke but not coronary heart disease.(10) Another study of 4,348 patients with type 2 diabetes found an inverse relationship between serum magnesium and incident HF and atrial fibrillation, but not with MACE.(9) Our nationwide ambulatory cohort of 1.2 million patients with type 2 diabetes builds upon these results by not only showing the associated risks of hypomagnesemia with MACE and death but also the association with improved outcomes with prescribed magnesium.

Potential mechanisms that may relate low magnesium to MACE and death in patients with type 2 diabetes include worsening of CVD risk factors,(11–13) oxidative stress,(6) inflammation,(7) endothelial dysfunction, and atherosclerosis,(8) all of which may increase the risk of thromboembolic events such as AMI or stroke and worsening of HF.

Mg supplementation has been shown to improve insulin resistance and hyperglycemia,(34, 35) inflammation and endothelial dysfunction,(35, 36) as well as hypertension (37) in trials of 4-26 weeks duration. Research also demonstrated that even short-term (<3 weeks) Mg supplementation can reduce the risk of ventricular arrhythmias in heart failure.(38–40) In our study, the median duration of magnesium prescription was 90 days (interquartile range [IQR], 30-176 days), making it biologically plausible that prescribed magnesium corrects hypomagnesemia, stabilizes cardiac function, and improves cardiovascular outcomes.

### 4.2 Hypermagnesemia and Cardiovascular Outcomes

Hypermagnesemia was also associated with a higher one-year risk of MACE and death in our ambulatory cohort of patients with type 2 diabetes. Similar associations between hypermagnesemia and an increased risk of adverse cardiovascular outcomes were observed in patients with HF in a systematical meta-analysis (41) and a prospective study (n = 500) of hospitalized patients.(42) Potential mechanisms by which hypermagnesemia can cause harm include muscular weakness (including respiratory muscles) and discoordination, profound fatigue, hypotension, bradycardia, and other cardiac dysrhythmias, as well as confusion and altered mental status.(43)

### 4.3 Clinical implications

These findings have significant clinical implications. Monitoring serum magnesium levels is currently not a standard practice in the outpatient management of patients with type 2 diabetes. Our results suggest that this should be considered in certain patients at risk, as both low and high magnesium levels were associated with adverse outcomes. Examples of such patients from our results may include those with cardiac or neurologic disorders, alcohol abuse, liver disease, abnormal kidney function, and those on medications that may alter magnesium excretion, such as thiazide diuretics, PPIs, or metformin. The subgroup analyses of patients with hypomagnesemia indicated that the impact of prescribed magnesium was more pronounced in patients taking PPIs or thiazides. These findings also highlight opportunities for practice improvement since prescribed magnesium was observed in <10% of patients with hypomagnesemia. The finding that prescribed magnesium is not associated with improved outcomes in patients with normomagnesemia is novel. It suggests that the body’s magnesium levels are maintained within a narrow normal range for optimal function.(2, 44, 45)

### 4.4 Limitations

This study has several limitations. First, the study population was predominantly male, which may limit the generalizability of our findings to females. However, our cohort included 11,705 female patients in the hypomagnesemia group and 44,640 in the normomagnesemia group, substantial numbers compared with other studies. Second, while we combined VHA and Medicare data for outcome ascertainment, patients seeking care from other payers may have been missed, leading to potential misclassification bias and underestimation of the associations. Third, restricting the sample to patients with serum magnesium levels may introduces a potential selection bias, possibly favoring the inclusion of a higher-risk population with comorbidities or conditions necessitating magnesium testing, as such, our results should be interpreted accordingly. However, it is important to note that our study cohort represents over half of the VHA users with type 2 diabetes, suggesting that we captured a substantial proportion of the at-risk population. Fourth, despite our new-user design, propensity-matching, sensitivity, and subgroup analyses to minimize confounding, residual confounding may still exist, given the study’s observational nature. This limitation can be overcome by conducting a large-scale RCT in the future. Fifth, our replacement study was limited by evaluating a binary treatment strategy (magnesium replacement vs. no replacement), without accounting for the potential effects of varying magnesium dosages and treatment durations. Future research should explore these dose-response relationships. Furthermore, we did not conduct a follow-up analysis to examine changes in serum magnesium levels over time, which limits our ability to assess the long-term impact of magnesium replacement therapy on both serum magnesium levels and cardiovascular risk.

## 5. Conclusion

In summary, our study showed that in patients with type 2 diabetes, both hypomagnesemia and hypermagnesemia were associated with higher one-year risks of MACE and death compared to normomagnesemia. Prescribed oral magnesium was associated with reduced risks of MACE and death in hypomagnesemia but not in normomagnesemia.

## Data Availability

The data that support the findings of this study are derived from the VHA CDW.
Restrictions apply to the availability of these data, which were used under the data-user-
agreement for this study. Data are available to VHA researchers upon adequate
approval.

## Data Sharing Statement

The data that support the findings of this study are derived from the VHA CDW. Restrictions apply to the availability of these data, which were used under the data-user-agreement for this study. Data are available to VHA researchers upon adequate approval.

## Author Contributions

Y. Y. collected data, conducted the analyses, and wrote the first draft of the manuscript. W.W., Q. Z., and A.A. conceptualized the project, acquired funding, and provided data interpretation. Y.C. and A.R.Z contributed to the study design and provided statistical input. All authors contributed to the writing and reviewed and edited the manuscript. All authors approved the final version of the manuscript.

