## Supplementary material for "Serum Magnesium, Prescribed Magnesium Replacement and Cardiovascular Events in Adults with Type 2 Diabetes: A National Cohort Study in U.S. Veterans": online supplements

**eFigure 1. Assembly of the Study Cohort.**

*Abbreviation: Mg = magnesium; T2D = type-2 diabetes*

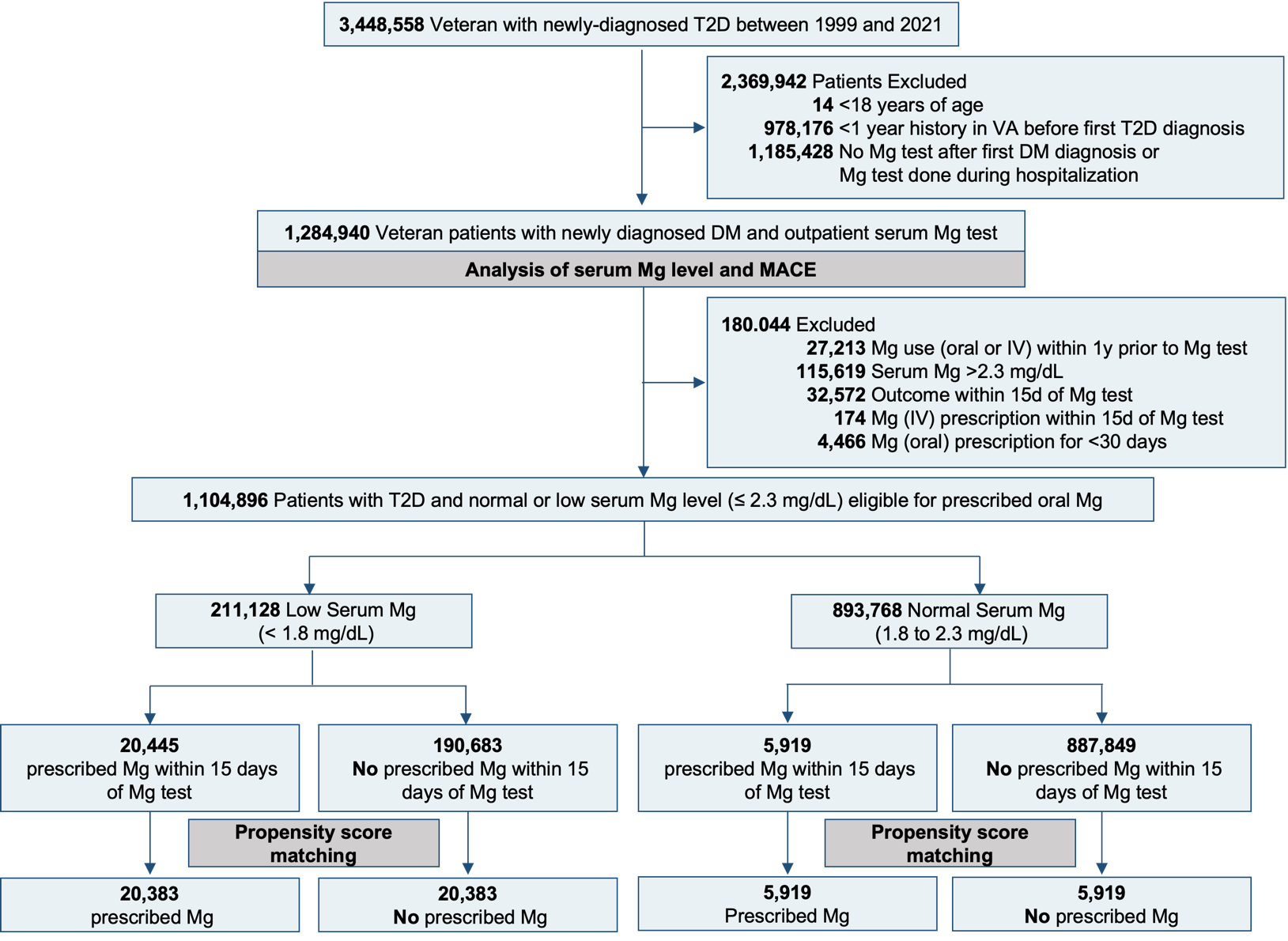

**eFigure 2: Love Plots Displaying the Absolute Standardized Difference of 64 Baseline Characteristics between Patients with Type-2 Diabetes and A) Hypomagnesemia, or B) Normomagnesemia, Who Were and Were Not Initiated on Prescribed Magnesium Before and After Propensity Score Matching**

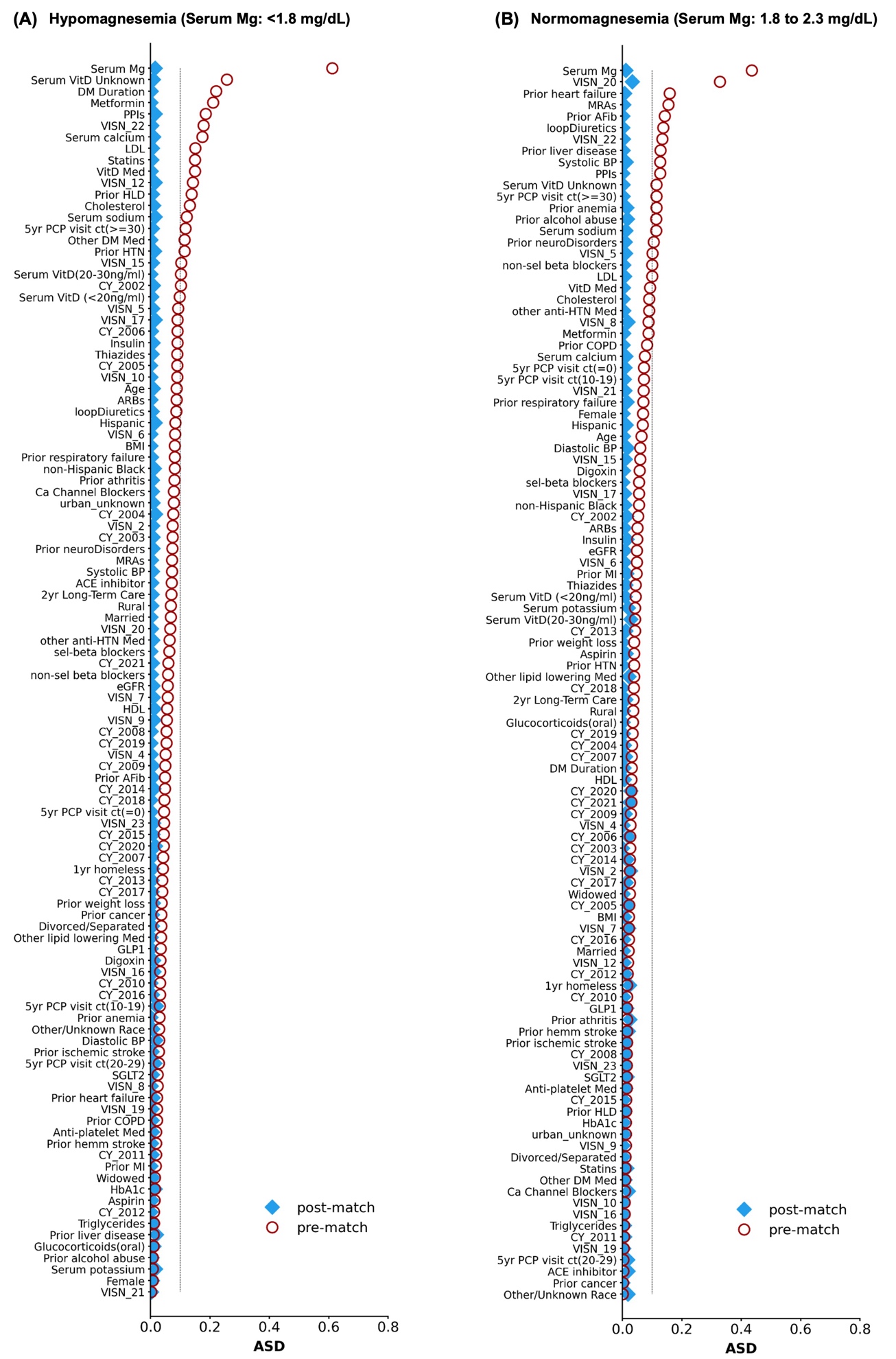

**eFigure 3: One-Year Kaplan-Meier Curves for Survival by Initiation of Prescribed Magnesium in Propensity Score-Matched Cohorts of Patients with Type 2 Diabetes and A) Hypomagnesemia, or B) Normomagnesemia**

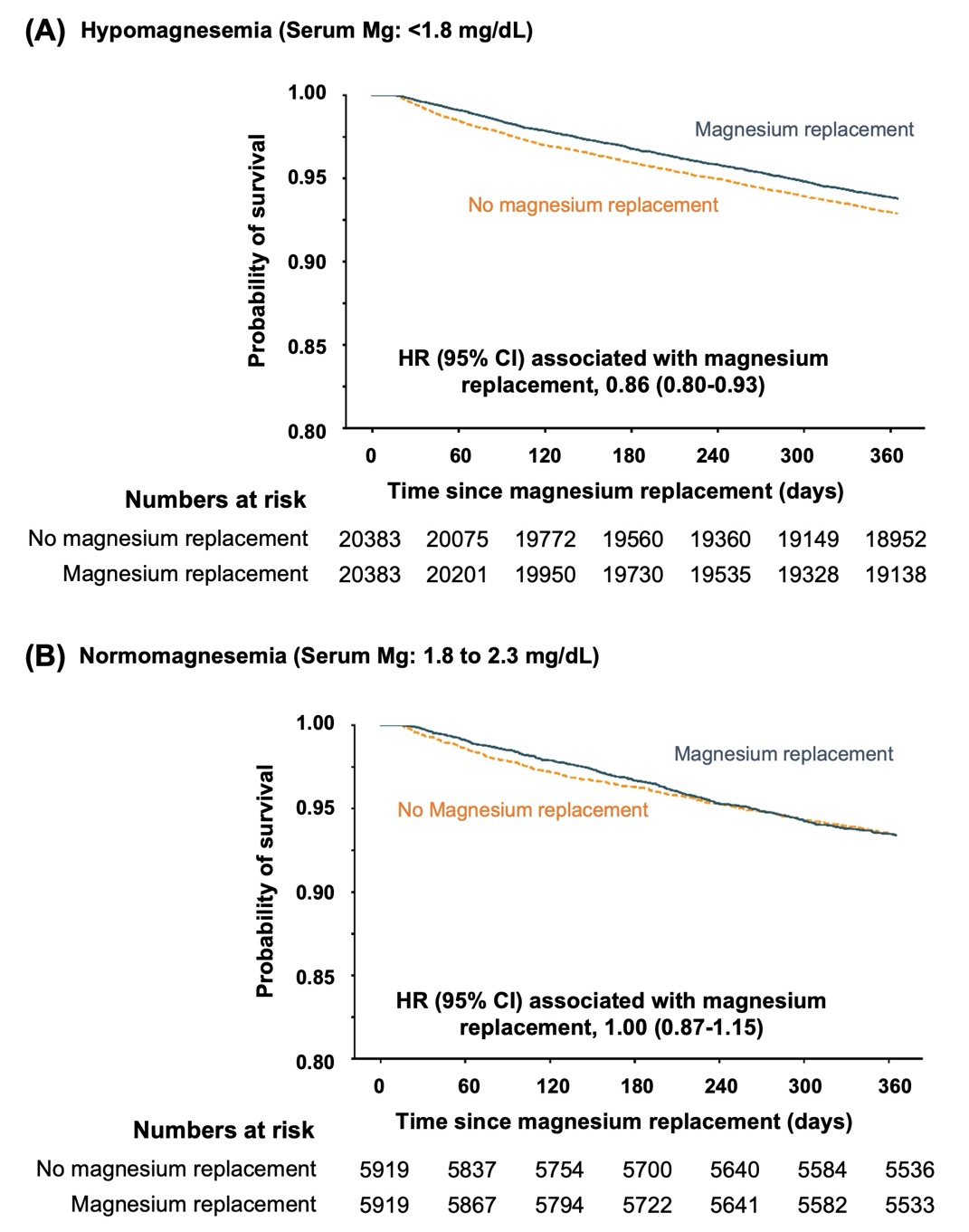

**eFigure 4. Hazard Ratios for the Association of Prescribed Magnesium and Time to MACE Within One Year in Subgroup-Specific Propensity-Matched Cohorts of Patients With Type 2 Diabetes and Hypomagnesemia**

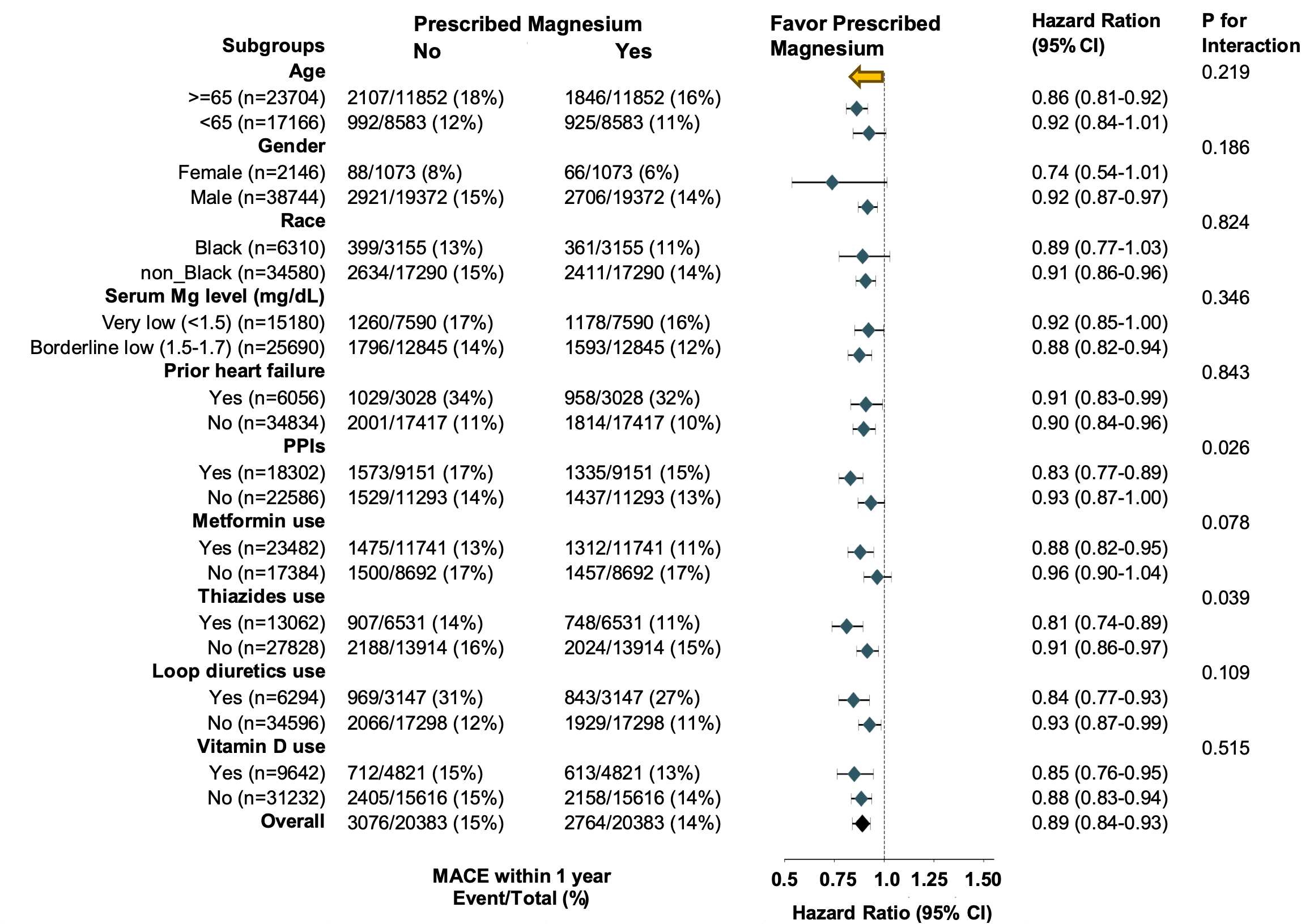

**eFigure 5. Hazard Ratios for the Association of Prescribed Magnesium and Time to MACE Within One Year in Subgroup-Specific Propensity-Matched Cohorts of Patients With Type 2 Diabetes and Normomagnesemia**

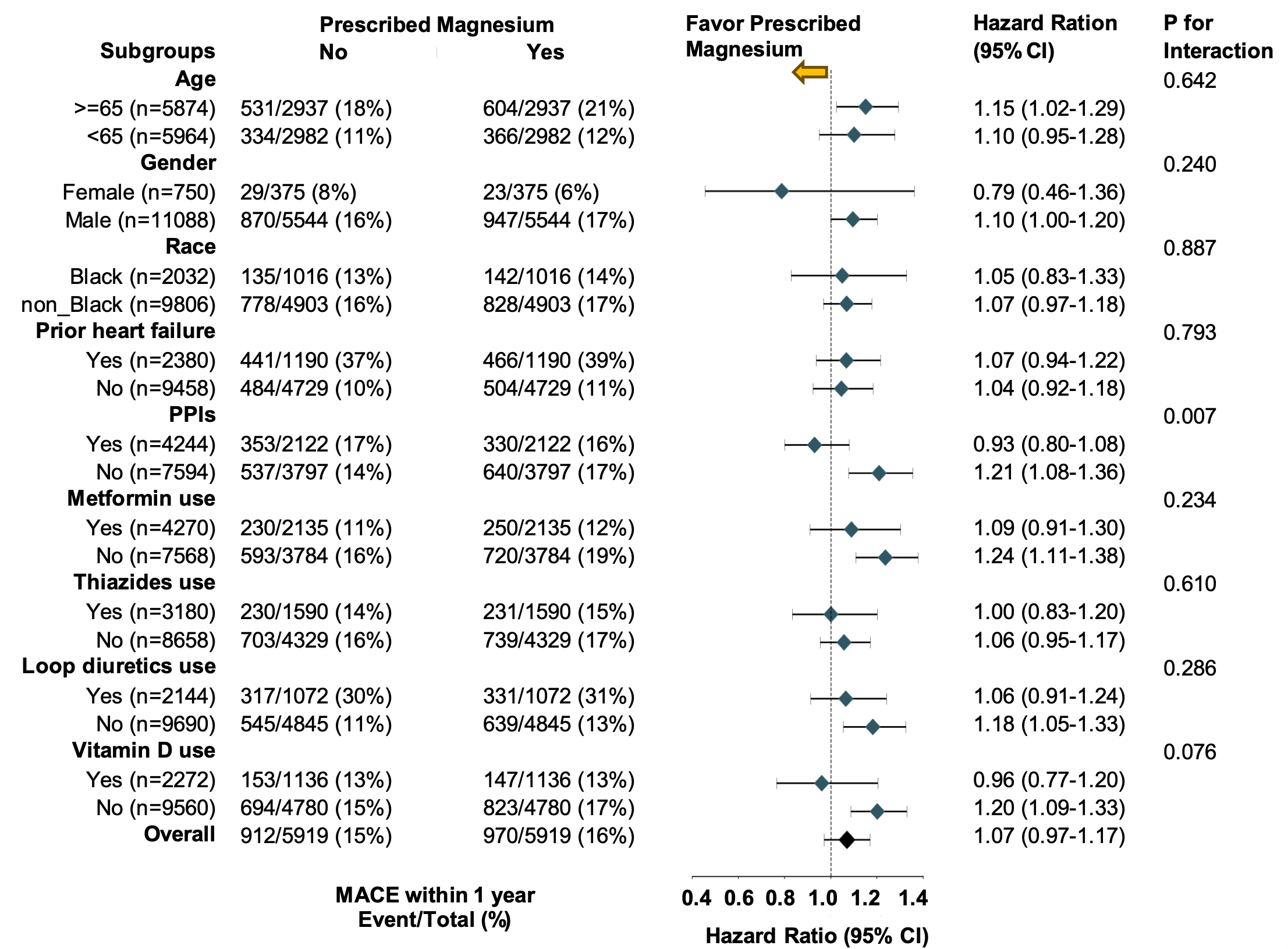

**eTable1: Diagnosis Codes**

| **Condition** | **ICD-9CM** | **ICD-10CM** |
| --- | --- | --- |
| Type 2 Diabetes | 250.X,357.2X,362.0X,366.41,648.0X | E11.x, O24.1x |
| AMI | 410.x, 412.x | I21.x, I22.x, I25.2x |
| Heart Failure | 428.x,398.91,402.01,402.11, | I50.x, I11.0x, I13.0x, I13.2x |
|  | 402.91,404.01,404.03,404.11, |  |
|  | 404.13,404.91,404.98 |  |
| Ischemic stroke | 436.x, 438.x,433.01,433.11, | G46.x, I63.x, I69.3x, I69.8x, I69.9x |
|  | 433.21,433.31,433.81,433.91, |  |
|  | 434.01,434.11,434.91 |  |
| Hemorrhagic stroke | 430.x, 431.x, 432.x | I60, I61, I62, I69.0, I69.1, I69.2 |
| Atrial Fibrillation | 427.31 | I48.0x, I48.1x, I48.2x, I48.91 |
| Hyperlipidemia | 272.0x,272.1x,272.2x,272.3x,272.4x | E78.0x, E78.1x, E78.2x, E78.3x, |
|  |  | E78.4x, E78.5x |
| Hypertension | 401.x | I10.x, I16.x |
| Alcohol abuse | 303.x,305.0x | F10.x |
| Anemia | 280.x-285.x | D50.x-D53.x, D55.x-D64.x |
| Arthritis | 715.x, 714.x,716.2x,716.3x, | M05.x-M08.x, M10.x, M13.x, M1A.x, |
|  | 716.5x,716.6x | M15.x-M19.x |
| Cancer | 140.x -208.x, 230.x-239.x | C00.x-D09.x D37.x-D49.x |
| COPD | 491.x, 492.x, 496.x | J41.x, J42.x, J43.x, J44.x |
| Liver Disease | 570.x-573.x | B15.x-B19.x, K70.x-K77.x |
| Neuro Disorders | 320.x-327.x, 330.x-337.x, | G00.x-G14.x, G21.x-G26.x,G30.x-G32.x, |
|  | 339.x-341.x, 345.x-349.x | G35.x-G37.x, G40, G43, G44, G47 |
| Respiratory Failure | 799.1,518.51,518.53,518.81,518.84 | J96, J80, R09.2, J95.82 |
| Weight Loss | 260.x-263.x,799.4,783.2,783.7 | E40.x-E46.x, R64.x, R63.4x |

**eTable2: Medication List**

| **Medication Group** | **Drug Name (generic)** |
| --- | --- |
| Diabetes Medications |  |
| Insulin | Insulin |
| Metformin | Metformin |
| GLP-1 receptor agonists | Albiglutide, Dulaglutide, Liraglutide, Lixisenatide, Semaglutide |
| SGLT-2 Inhibitors | Canaglifozin, Dapagliflozin, Empagliflozin |
| Others | Thiazolidinediones: Pioglitazone, Rosiglitazone, Troglitazone |
| Cardiovascular Medications | |
| ACEIs | Benazepril, Captopril, Enalapril, Enalaprilat, Fosinopril, Lisinopril, Moexipril, Perindopril, Quinapril, Ramipril, Trandolapril |
| ARBs | Azilsartan,Candesartan,Eprosartan,Irbesartan,Losartan, Olmesartan,Telmisartan,Valsartan |
| Other anti-hypertensive medications | Aliskiren, Amiloride, Chlorpromazine, Chlorthalidone, Clonidine, Doxazosin, Eplerenone, Hydralazine, Methyldopa, Methyldopate, Nitroprusside, Phenoxybenzamine, Prazosin, Spironolactone, Terazosin, Tolazoline, Triamterene |
| Anti-platelets | Abciximab, Cangrelor, Cilostazol, Clopidogrel, Dipyridamole, Eptifibatide, Prasugrel, Ticagrelor, Ticlopidine, Tirofiban, Asprin |
| Selective beta blockers | Acebutolol, Atenolol, Betaxolol, Bisoprolol, Esmolol, Metoprolol, Nebivolol |
| Non-selective beta blockers | Carteolol, Carvedilol, Labetalol, Levobunolol, Metipranolol, Nadolol, Penbutolol, Pindolol, Propranolol, Timolol |
| Calcium Channel Blockers | Amlodipine, Clevidipine, Diltiazem, Felodipine, Isradipine, Levamlodipine, Nicardipine, Nifedipine, Nimodipine, Verapamil |
| Digoxin and other Inotropes | Amrinone Inj, Digoxin, Dobutamine Inj, Dopamine Inj, Disopyramide, Epinephrine, Ethylnorepinephrine Inj, Flecainide, Inamrinone Inj, Isoproterenol Inj, Milrinone,Norepinephrine Inj, Procainamide, Quinidine, Sunitinib, Theophylline |
| Loop Diuretics | Bumetanide, Ethacrynic Acid, Furosemide |
| MRAs | Eplerenone, Finerenone, Spironolactone |
| Other non-statin lipid lowering | Alirocumab, Cholestyramine, Clofibrate, Colesevelam, Colestipol, Evolocumab, Ezetimibe, Fenofibrate, Fenofibric Acid, Gemfibrozil, Inclisiran, Lomitapide, Niacin |
| Statins | Atorvastatin, Cerivastatin, Fluvastatin, Lovastatin, Pitavastatin, Pravastatin, Rosuvastatin, Simvastatin |
| Thiazides | Chlorothiazide, Hydrochlorothiazide, Indapamide, Methyclothiazide, Metolazone |
| Select Non-Cardiovascular Medications | |
| Vitamin D | Ergocalciferol (Vitamin D2), Cholecalciferol (Vitamin D3) |
| Glucocorticoids | Cortisone, Dexamethasone, Fludrocortisone, Hydrocortisone, Methylprednisolone, Prednisolone, Prednisone |
| PPIs | Dexlansoprazole, Esomeprazole, Omeprazole, Panteprazole, Rabeprazole |

**eTable 3: Listing of Continuous Variables with Missing Data**

|  | **Hypomagnesemia**  **(Serum Magnesium: <1.8 mg/dL)** | | **Normomagnesemia**  **(Serum Magnesium: 1.8 to 2.3 mg/dL)** | |
| --- | --- | --- | --- | --- |
|  | **Prescribed Magnesium** | | **Prescribed Magnesium** | |
|  | **No** | **Yes** | **No** | **Yes** |
| **Variables^a^, N (%)** | **(n= 190,683)** | **(n= 20,445)** | **(n= 887,849)** | **(n= 5,919)** |
| HbA1c | 22954 (12%) | 1236 (6%) | 116207 (13%) | 705 (12%) |
| BMI | 11165 (6%) | 1036 (5%) | 52955 (6%) | 254 (4%) |
| Serum Sodium | 3556 (2%) | 301 (1%) | 17703 (2%) | 128 (2%) |
| Serum Potassium | 2949 (2%) | 256 (1%) | 16876 (2%) | 198 (3%) |
| Serum Calcium | 10683 (6%) | 1440 (7%) | 53396 (6%) | 401 (7%) |
| Systolic BP | 5132 (3%) | 552 (3%) | 25172 (3%) | 104 (2%) |
| Diastolic BP | 5132 (3%) | 552 (3%) | 25172 (3%) | 104 (2%) |
| LDL cholesterol | 31814 (17%) | 2231 (11%) | 131389 (15%) | 894 (15%) |
| Triglycerides | 29631 (16%) | 2274 (11%) | 125421 (14%) | 821 (14%) |
| Cholesterol | 28370 (15%) | 1907 (9%) | 121090 (14%) | 778 (13%) |
| HDL cholesterol | 26095 (14%) | 1837 (9%) | 111487 (13%) | 697 (12%) |
| eGFR | 5150 (3%) | 537 (3%) | 32938 (4%) | 178 (3%) |
| *^a^Lab test and vital sign data assessed within one year before the index date; if there is no data available, then consider it as missing. Missing data were imputed by single imputation using a fitted general linear model on age, sex, race and ethnicity* | | | | |

**eTable 4. Baseline Characteristics of Patients with Type-2 Diabetes and Normomagnesemia Initiated on Prescribed Oral Magnesium, Before and After Propensity Score Matching**

| **Characteristic, N (%)** | **Before matching (n=893,768)** | | | **After matching (n=11,838)** | |
| --- | --- | --- | --- | --- | --- |
|  | **Prescribed Magnesium** | | **Prescribed Magnesium** | | |
|  | **No** | **Yes** | **No** | | **Yes** |
|  | **(n= 887,849)** | **(n= 5,919)** | **(n= 5,919)** | | **(n= 5,919)** |
| **Age, Mean (SD), y** | 65.4 (11.8) | 64.7 (11.3) | 64.7 (11.7) | | 64.7 (11.3) |
| **Gender** |  |  |  | |  |
| Male | 845672 (95) | 5544 (94) | 5542 (94) | | 5544 (94) |
| Female | 42177 (5) | 375 (6) | 377 (6) | | 375 (6) |
| **Diabetes Duration, Mean (SD), y** | 3.6 (4.0) | 3.7 (4.2) | 3.7 (4.1) | | 3.7 (4.2) |
| **Serum Magnesium, Mean (SD), mg/dL** | 2.03 (0.15) | 1.96 (0.16)^e^ | 1.96 (0.14) | | 1.96 (0.16) |
| **Race** |  |  |  | |  |
| Hispanic | 52710 (6) | 262 (4) | 277 (5) | | 262 (4) |
| non-Hispanic White | 574711 (65) | 4046 (68) | 4016 (68) | | 4046 (68) |
| non-Hispanic Black | 171271 (19) | 1016 (17) | 998 (17) | | 1016 (17) |
| Other/Unknown | 89157 (10) | 595 (10) | 628 (11) | | 595 (10) |
| **Marital Status** |  |  |  | |  |
| Single | 86810 (10) | 532 (9) | 530 (9) | | 532 (9) |
| Divorced/Separated | 243638 (27) | 1652 (28) | 1660 (28) | | 1652 (28) |
| Married | 480844 (54) | 3264 (55) | 3255 (55) | | 3264 (55) |
| Widowed | 76557 (9) | 471 (8) | 474 (8) | | 471 (8) |
| **Cormobidities^a^** |  |  |  | |  |
| Heart Failure | 125568 (14) | 1190 (20)^e^ | 1173 (20) | | 1190 (20) |
| AMI | 95152 (11) | 724 (12) | 697 (12) | | 724 (12) |
| Ischemic stroke | 79737 (9) | 507 (9) | 524 (9) | | 507 (9) |
| Hemorrhagic stroke | 7283 (1) | 41 (1) | 32 (1) | | 41 (1) |
| Atrial Fibrillation | 105248 (12) | 996 (17)^e^ | 998 (17) | | 996 (17) |
| Hyperlipidemia | 704870 (79) | 4671 (79) | 4653 (79) | | 4671 (79) |
| Hypertension | 758041 (85) | 5133 (87) | 5131 (87) | | 5133 (87) |
| Anemia | 203332 (23) | 1649 (28)^e^ | 1612 (27) | | 1649 (28) |
| Alcohol abuse | 160005 (18) | 1337 (23)^e^ | 1378 (23) | | 1337 (23) |
| Liver Disease | 93063 (10) | 871 (15)^e^ | 874 (15) | | 871 (15) |
| Respiratory Failure | 30089 (3) | 282 (5) | 263 (4) | | 282 (5) |
| COPD | 226104 (25) | 1723 (29) | 1729 (29) | | 1723 (29) |
| Cancer | 269316 (30) | 1799 (30) | 1797 (30) | | 1799 (30) |
| Neuro Disorders | 312089 (35) | 2380 (40)^e^ | 2354 (40) | | 2380 (40) |
| Weight Loss | 62253 (7) | 476 (8) | 476 (8) | | 476 (8) |
| Arthritis | 411169 (46) | 2785 (47) | 2712 (46) | | 2785 (47) |
| Gagne Comorbidity Score^b^ | 1.9 (2.4) | 2.3 (2.5)^e^ | 2.3 (2.6) | | 2.3 (2.5) |
| **Comedications^c^** |  |  |  | |  |
| Diabetes Medications |  |  |  | |  |
| Insulin | 108472 (12) | 822 (14)^e^ | 853 (14) | | 822 (14) |
| Metformin | 283582 (32) | 2135 (36) | 2146 (36) | | 2135 (36) |
| GLP1 | 5780 (1) | 46 (1) | 39 (1) | | 46 (1) |
| SGLT2 inhibitors | 8522 (1) | 65 (1) | 75 (1) | | 65 (1) |
| Other Diabetes Medication | 185068 (21) | 1213 (20) | 1233 (21) | | 1213 (20) |
| Other Medications |  |  |  | |  |
| Thiazides | 221249 (25) | 1590 (27) | 1561 (26) | | 1590 (27) |
| Loop Diuretics | 116892 (13) | 1073 (18)^e^ | 1072 (18) | | 1073 (18) |
| PPIs | 265782 (30) | 2122 (36)^e^ | 2120 (36) | | 2122 (36) |
| Vitamin D | 139702 (16) | 1139 (19) | 1133 (19) | | 1139 (19) |
| ACEIs | 375259 (42) | 2494 (42) | 2547 (43) | | 2494 (42) |
| ARBs | 97089 (11) | 743 (13) | 732 (12) | | 743 (13) |
| Other anti-hypertension | 195342 (22) | 1529 (26) | 1520 (26) | | 1529 (26) |
| Selected beta blockers | 257551 (29) | 1868 (32) | 1866 (32) | | 1868 (32) |
| Non-selected beta blockers | 81962 (9) | 730 (12)^e^ | 722 (12) | | 730 (12) |
| Digoxin or Other Inotropes | 36528 (4) | 316 (5) | 313 (5) | | 316 (5) |
| Aspirin | 180812 (20) | 1299 (22) | 1328 (22) | | 1299 (22) |
| Anti-platelet | 65983 (7) | 419 (7) | 403 (7) | | 419 (7) |
| Glucocorticoids | 86120 (10) | 637 (11) | 634 (11) | | 637 (11) |
| Statins | 495418 (56) | 3274 (55) | 3314 (56) | | 3274 (55) |
| Other non-statin lipid lowering | 85265 (10) | 503 (8) | 469 (8) | | 503 (8) |
| Calcium Channel Blocker | 226774 (26) | 1532 (26) | 1483 (25) | | 1532 (26) |
| MRAs | 27328 (3) | 376 (6)^e^ | 370 (6) | | 376 (6) |
| **Health examination data^d^** |  |  |  | |  |
| HbA1c, Mean (SD), % | 7.0 (1.6) | 7.1 (1.7) | 7.1 (1.7) | | 7.1 (1.7) |
| BMI, Mean (SD), kg/m2 | 30.8 (6.6) | 31.0 (6.9) | 30.9 (6.7) | | 31.0 (6.9) |
| Systolic BP, Mean (SD), mmHg | 134.8 (19.0) | 132.3 (19.0)^e^ | 132.5 (19.0) | | 132.3 (19.0) |
| Diastolic BP, Mean (SD), mmHg) | 76.5 (11.7) | 75.8 (11.7) | 76.0 (11.8) | | 75.8 (11.7) |
| eGFR, Mean (SD), mL/min/1.73m2 | 75.0 (23.3) | 73.8 (24.7) | 73.9 (24.1) | | 73.8 (24.7) |
| Serum Vitamin D, Mean (SD), ng/mL |  |  |  | |  |
| <20 | 54513 (6) | 428 (7) | 414 (7) | | 428 (7) |
| 20-30 | 84101 (9) | 636 (11) | 687 (12) | | 636 (11) |
| 30-100 | 127068 (14) | 1025 (17) | 993 (17) | | 1025 (17) |
| Unknown | 622167 (70) | 3830 (65)^e^ | *3825 (65)* | | 3830 (65) |
| Serum Sodium, Mean (SD), mEq/L | 138.4 (3.4) | 138.0 (3.8)^e^ | 138.0 (3.5) | | 138.0 (3.8) |
| Serum Potassium, Mean (SD), mEq/L | 4.2 (0.5) | 4.2 (0.6) | 4.2 (0.5) | | 4.2 (0.6) |
| Serum Calcium, Mean (SD), mg/dL | 9.2 (0.6) | 9.2 (0.8) | 9.2 (0.7) | | 9.2 (0.8) |
| LDL cholesterol, Mean (SD), mg/dL | 95.1 (38.8) | 91.0 (35.5)^e^ | 91.2 (35.2) | | 91.0 (35.5) |
| Triglycerides, Mean (SD), mg/dL | 170.6 (114.7) | 169.4 (117.7) | 168.4 (114.8) | | 169.4 (117.7) |
| Cholesterol, Mean (SD), mg/dL | 168.3 (45.2) | 164.0 (46.5) | 163.8 (43.3) | | 164.0 (46.5) |
| HDL cholesterol, mg/dL | 41.7 (12.8) | 42.2 (13.8) | 42.3 (13.6) | | 42.2 (13.8) |
| **Patient Residence** |  |  |  | |  |
| Rural | 116516 (13) | 850 (14) | 856 (14) | | 850 (14) |
| Urban | 421988 (48) | 2772 (47) | 2751 (46) | | 2772 (47) |
| Unknown | 349345 (39) | 2297 (39) | *2312 (39)* | | 2297 (39) |
| **PCP visits in the past five year** |  |  |  | |  |
| 0 | 30911 (3) | 291 (5) | 281 (5) | | 291 (5) |
| 1-9 | 196916 (22) | 1116 (19) | 1147 (19) | | 1116 (19) |
| 10-19 | 247240 (28) | 1462 (25) | 1477 (25) | | 1462 (25) |
| 20-29 | 181158 (20) | 1200 (20) | 1159 (20) | | 1200 (20) |
| >=30 | 231624 (26) | 1850 (31)^e^ | 1855 (31) | | 1850 (31) |
| **Homeless in past year** | 42453 (5) | 303 (5) | 334 (6) | | 303 (5) |
| **Long-Term Care in past two year** | 13077 (1) | 63 (1) | 70 (1) | | 63 (1) |

*^a^Diagnoses assessed before the index date.*

*^b^A weighted combined comorbidity score.*

*^c^Medication prescription assessed one year before the index date*

*^d^Latest examination data assessed within one year before the index date*

*^e^With more than a 10% absolute standardized difference compared to patients with no prescribed magnesium before propensity score matching*

**eTable 5. Outcomes During One Year of Follow-Up for Patients With Type-2 Diabetes and Hypomagnesemia (Top Panel) and Diabetes and Normomagnesemia (Bottom Panel) by Prescribed Magnesium**

|  | **Before Propensity Score Matching** | | | **After Propensity Score Matching** | |  |
| --- | --- | --- | --- | --- | --- | --- |
|  | **Events (%) by prescribed magnesium in patient with hypomagnesemia** | | **Adjusted HR^a^ (95% CI)** | **Events (%) by prescribed magnesium in patient with hypomagnesemia** | | **HR^b^ (95% CI)** |
|  | **No (n= 190,683)** | **Yes (n=20,445)** |  | **No (n= 20,383)** | **Yes (n=20,383)** |  |
| MACE | 29846 (15.7%) | 2772 (13.6%) | 0.93 (0.89-0.96) | 3076 (15.1%) | 2764 (13.6%) | 0.89 (0.84-0.93) |
| All-cause mortality | 14410 (7.6%) | 1267 (6.2%) | 0.93 (0.88-0.99) | 1452 (7.1%) | 1263 (6.2%) | 0.86 (0.80-0.93) |
|  | **Events (%) by prescribed magnesium in patient with normomagnesemia** | | **Adjusted HR^a^ (95% CI)** | **Events (%) by prescribed magnesium in patient with normomagnesemia** | | **HR^b^ (95% CI)** |
|  | **No (n=887,849)** | **Yes (n=5,919)** |  | **No (n=5,919)** | **Yes (n=5,919)** |  |
| MACE | 115632 (13.0%) | 970 (16.4%) | 1.14 (1.07-1.21) | 912 (15.4%) | 970 (16.4%) | 1.07 (0.97-1.17) |
| All-cause mortality | 48537 (5.5%) | 391 (6.6%) | 1.11 (1.01-1.23) | 389 (6.6%) | 391 (6.6%) | 1.00 (0.87-1.15) |
| *^a^The adjusted hazard ratios are associated with prescribed-oral-magnesium when compared with no prescribed oral magnesium, assessed using a multivariable Cox regression model, adjusted for 64 covariates. These include demographic information (age, sex, race, etc.), clinical characteristics (comorbidities, comedications, health examination data, etc.), healthcare utilization metrics, geographic information, and index year.* | | | | | | |
| *^b^The hazard ratios are associated with prescribed-oral-magnesium when compared with no Prescribed-oral-magnesium* | | | | |  |  |
